# EEG-Based Identification of Adolescent Non-Suicidal Self-Injury and Neurophysiological Interpretation Using an Explainable Deep Learning Framework

**DOI:** 10.64898/2026.06.23.26356351

**Authors:** Tianqi Liu, Xinyi Liu, Yihang Bao, Wenhao Li, Guan Ning Lin

## Abstract

Non-suicidal self-injury (NSSI) among adolescents is a prevalent mental health problem and an important indicator of potential suicide risk. Early objective identification and neural mechanism analysis are therefore crucial for clinical screening and intervention. Traditional assessments mainly rely on self-report scales and clinical interviews, which are vulnerable to subjective bias, clinical experience, and missed diagnosis. Electroencephalography (EEG), with its non-invasive, low-cost, and high-temporal-resolution characteristics, provides a promising physiological basis for identifying NSSI-related neural abnormalities. However, EEG-based intelligent recognition of adolescent NSSI remains limited, and existing studies often emphasize classification performance while lacking systematic neurophysiological interpretation. To address these issues, this study proposes CGA-NSSI, a lightweight deep learning framework for adolescent NSSI recognition. The model integrates a one-dimensional convolutional neural network, bidirectional gated recurrent unit, and multi-head self-attention mechanism to extract local spatiotemporal EEG features, model long-range temporal dependencies, and focus on key pathology-related time segments and channels. A standardized preprocessing pipeline, together with Mixup augmentation and Focal Loss, is further used to alleviate sample imbalance and improve robustness in small clinical EEG datasets. Experiments on a real-world adolescent clinical EEG dataset show that CGA-NSSI can effectively identify NSSI-related EEG patterns under imbalanced sample conditions. Interpretability and functional connectivity analyses further reveal prefrontal-centered cross-regional network reorganization, excessive static functional coupling, reduced dynamic connectivity fluctuations, and increased abnormal state occupancy. These findings suggest that CGA-NSSI not only improves objective NSSI recognition but also provides neurophysiological evidence for understanding adolescent self-injury.

## 1. Introduction

Non-suicidal self-injury (NSSI) is defined as the deliberate, repetitive infliction of bodily harm in the absence of suicidal intent [1]. It has been formally recognized as an independent condition in the Diagnostic and Statistical Manual of Mental Disorders, Fifth Edition (DSM-5) as a condition for further study [2]. Over the past decade, the prevalence of NSSI among adolescents and young adults has risen sharply, making it a growing public health concern [3, 4]. In China, the overall prevalence of NSSI among adolescents is estimated at approximately 29.1% [5]. Beyond its immediate harm, NSSI is closely associated with impaired psychosocial functioning [6] and is a significant predictor of future suicidal behavior [7]. Accordingly, elucidating the mechanisms underlying NSSI and developing reliable early-identification tools are matters of considerable clinical urgency.

Electroencephalography (EEG) provides a direct window into the neuronal dynamics originating from the central nervous system [8]. As a cost-effective, non-invasive modality with high temporal resolution, EEG is highly valuable for detecting abnormal brain activity [9, 10]. EEG has been widely applied across various disease domains, particularly in psychiatric and neurological conditions, including Alzheimer’s diseaseA [11], affective disorders [12], depression [13], autism spectrum disorder (ASD) [14], and schizophrenia [15]. These applications underscore the versatility and importance of EEG in understanding and intervening in neurological and psychiatric disorders. However, research leveraging EEG combined with deep learning for the diagnosis of adolescent self-injurious behavior remains notably scarce.

Existing research on adolescent self-injury has largely concentrated on elucidating neurophysiological mechanisms [16–21]. Converging evidence indicates that both NSSI and suicidal behavior involve multi-regional brain network abnormalities, primarily manifesting as frontolimbic dysfunction— including dysregulation of the prefrontal cortex (emotion regulation), the amygdala (emotional reactivity), and the anterior cingulate cortex (impulse control), with hyperactivation and reduced connectivity in regions such as the dorsolateral prefrontal cortex (dlPFC). Beyond mechanistic inquiry, the field has progressively explored intelligent diagnostic approaches, with studies employing machine learning and regression models on emotional rating scales and neuroimaging data [22–25]. More recently, EEG-based deep learning models have been developed and validated with encouraging recognition performance [26]. Nevertheless, critical shortcomings remain: most models adopt a single-architecture design that struggles to simultaneously capture local EEG features and long-range temporal dependencies; and models generally lack interpretability, failing to connect learned representations to the established frontolimbic neurophysiological mechanisms of NSSI.

To address these limitations, we propose CGA-NSSI, a 1D CNN–BiGRU–Multi-Head Self-Attention fusion model that exploits the high temporal resolution of EEG. Four uniform 1D convolutional blocks extract local spatiotemporal micro-features; a BiGRU captures long-range temporal dependencies; and multi-head self-attention adaptively focuses on pathologically relevant EEG time segments. A standardized preprocessing pipeline—encompassing multi-channel alignment, robust normalization, and multi-window dense sampling—is established, complemented by Mixup augmentation and Focal Loss to address sample imbalance and small-sample generalization. Building on high-accuracy NSSI identification, we further conduct extensive neurophysiological analyses that provide objective evidence for mechanistic interpretation of self-injurious behavior and form a corroborating link with the established frontolimbic dysfunction hypothesis. The main contributions of this work are as follows:

- We develop CGA-NSSI, a lightweight network for imbalanced NSSI EEG, integrating 1D-CNN, BiGRU and multi-head attention to extract hierarchical spatiotemporal features.
- We validate our model on clinical EEG data and confirm its superior classification performance for clinical NSSI screening against mainstream benchmarks.
- We implement model interpretation and multi-scale brain connectivity analyses to build a spatial-temporal-behavior pathological framework and uncover core NSSI neuropathological changes: prefrontal over-coupling, fronto-temporo-parietal dysfunction and inflexible brain dynamics.

## 2 Related Work

### 2.1. Deep Learning-Based EEG Analysis in Psychiatric Disorders

Effective feature extraction from high-dimensional EEG signals is fundamental to elucidating brain function and enabling precise computational modeling. Conventional machine learning methods generally fail to capture the intrinsic nonlinear and dynamic properties of EEG [27]. Deep learning, by contrast, can automatically identify and represent information-rich spatiotemporal features, improving extraction accuracy and enabling dynamic adaptation to the non-stationary characteristics of neural activity [28]. Underpinned by standardized non-invasive EEG acquisition and analysis paradigms [29], researchers have progressively adopted sophisticated network architectures to probe dynamic pathological changes in the brain.

For instance, graph convolutional networks (GCN), Transformer architectures, and attention mechanisms have been widely applied to EEG feature extraction in neurological and psychiatric disorders. For Parkinson’s disease detection, Mancazzo et al. [11] employed a Transformer model with multi-head attention explanations, effectively achieving resting-state EEG-based Parkinson’s disease detection. In emotion recognition, Yan et al. [12] proposed a bridge graph attention-based graph convolutional network fused with a multi-scale Transformer, significantly improving the accuracy and robustness of EEG-based emotion recognition. For depression, Ying et al. [13] constructed a lightweight attention mechanism model based on brain functional connectivity, providing an efficient solution for EEG-based depression recognition. For ASD, Tseng et al. [14] fused local cortical activation features with global functional connectivity using a mobile EEG system and game-based stimuli, achieving more precise neural characterization of ASD. Notably, Guerra et al. [15] innovatively explored the diagnostic potential of large language models (LLMs) in EEG signal analysis, opening new frontiers for early objective detection of schizophrenia.

In summary, deep learning-based EEG methods have achieved considerable progress in the analysis of psychiatric disorders. However, research applying EEG combined with deep learning specifically to NSSI remains scarce.

### 2.2. EEG-Based Research on Adolescent NSSI

Despite the strong potential of deep learning in EEG-based psychiatric disorder analysis, the application of complex models to NSSI remains nascent. The trajectory of objective assessment research for adolescent NSSI traces a clear progression: from machine learning screening of basic physiological sensor data, toward advanced EEG spatiotemporal feature extraction, and finally to preliminary deep learning exploration.

In early explorations using objective physiological data for NSSI risk assessment, traditional machine learning techniques were first introduced. Marti-Puig et al. [22] explored physiological sensor data combined with machine learning classifiers to lay the groundwork for automated assessment of NSSI behavior in young adults. Building on this approach, Jiang et al. [23] further developed a specialized NSSI machine learning prediction model for rural Chinese junior high school students, validating the feasibility of data-driven objective assessment for early screening in specific adolescent populations.

To overcome the limitations of single-dimensional physiological signals and the lack of central nervous system mechanistic interpretation, researchers shifted focus to EEG with its high temporal resolution. Li et al. [24] combined EEG physiological features with psychological scales in adolescents with emotional disorders, establishing the indispensable value of EEG signals in comprehensive clinical objective assessment of NSSI.

To capture deeper dynamic spatiotemporal patterns in EEG signals, the research focus gradually shifted from basic EEG frequency-domain analysis to advanced dynamic feature extraction. Song et al. [25] extracted EEG microstate parameters and combined them with machine learning algorithms to successfully classify depressed adolescents with and without NSSI behavior, demonstrating that spatiotemporal dynamic changes in resting-state brain activity are effective neuroimaging indicators for distinguishing NSSI adolescents.

As frontier technologies converged, deep learning models for high-dimensional EEG data finally achieved breakthroughs in the NSSI domain. Tong et al. [26] pioneered the application of graph neural networks (GNN) to this field, constructing a GNN architecture based on a functional topology model for single-channel EEG data in an exploratory study with only three samples. Liang et al. [30] proposed NSSI-Net, a multi-concept generative adversarial network (GAN) framework operating under a semi-supervised mechanism to handle high-dimensional EEG data, alleviating the challenges of limited annotation and small sample sizes in clinical NSSI EEG data.

In summary, EEG-based adolescent NSSI analysis is steadily evolving from traditional machine learning toward deep learning integrating advanced spatiotemporal features. However, the lack of interpretability in existing models remains the primary bottleneck limiting clinical translation.

### 2.3. Neurophysiological Mechanisms in Adolescents with NSSI

Parallel to diagnostic modeling efforts, researchers have sought to elucidate the neurophysiological underpinnings of adolescent NSSI. Converging evidence from systematic reviews and neuroimaging studies indicates that adolescents with NSSI or suicidal behavior exhibit specific neural abnormalities at both structural and functional levels, providing an objective neurobiological basis for understanding impaired emotion regulation and impulsive behavior in this population [16, 17].

At the morphological and structural level, adolescents with NSSI commonly show tissue deficits in specific brain regions. Ando et al. [18] found that adolescents engaging in NSSI exhibited significant reductions in regional grey matter volume in certain brain areas, potentially directly undermining the underlying cognitive regulatory networks when facing intense negative emotions.

At the functional activity level, NSSI adolescents exhibit significantly abnormal neural responses when processing specific negative social-emotional stimuli. Groschwitz et al. [21] used fMRI to investigate brain activity changes in NSSI adolescents during social exclusion, revealing a specific neural processing pattern in response to negative social stimuli, which is considered an important functional driver of impulsive self-injurious behavior.

Beyond task-specific functional activation abnormalities, disrupted functional connectivity between core brain regions is central to understanding NSSI neural mechanisms. The amygdala, as the physiological hub for emotion generation and processing, plays a crucial role. Schreiner et al. [31] used multimodal neuroimaging to precisely localize and reveal significant abnormalities in amygdala functional connectivity in NSSI adolescents. This finding aligns with Santamarina-Perez et al. [20], who further confirmed that fronto-limbic connectivity is not only a core pathological feature of NSSI but also a key biological predictor of clinical improvement following psychotherapy.

As understanding of the neural mechanisms of NSSI deepens, research focus is extending toward mechanism-based interventions. Wu et al. [19] systematically summarized the complex neural mechanisms of NSSI and explored the potential of neuromodulation therapies in clinical intervention. These neurobiological findings enrich the objective assessment dimensions of NSSI and are driving treatment toward more precise targeted interventions.

## 3. Method

This section presents the CGA-NSSI framework in detail. The overall pipeline is illustrated in Fig. 1 and comprises three stages: (1) standardized EEG preprocessing, (2) hierarchical feature extraction via the cascaded 1D-CNN– BiGRU–MHSA network, and (3) global aggregation and binary classification.

**Figure 1.**
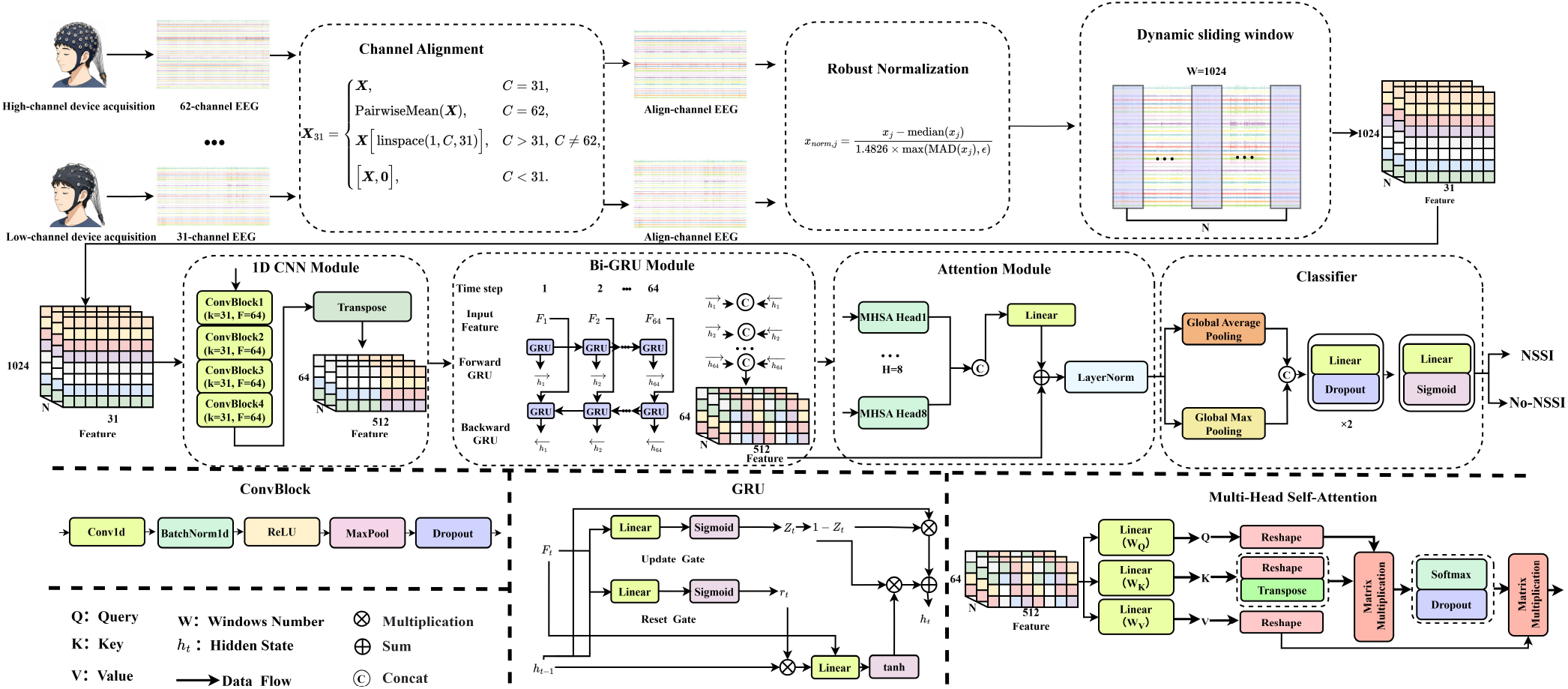
Overall framework of the proposed CGA-NSSI method. Raw EEG signals from heterogeneous devices are first standardized to 31 channels, robustly normalized, and segmented into fixed-length windows. The cascaded 1D-CNN–BiGRU– Multi-Head Self-Attention network then extracts hierarchical spatiotemporal features, followed by global pooling and an MLP classifier for end-to-end NSSI risk prediction.

### 3.1. Data Preprocessing

Raw EEG signals collected from heterogeneous acquisition devices exhibit substantial inter-subject variability in channel count, signal length, and artifact contamination (e.g., ocular and muscular artifacts). Before network ingestion, these sources of heterogeneity must be resolved. The preprocessing pipeline comprises three sequential steps: channel alignment, robust normalization, and dynamic sliding-window segmentation.

EEG data in our dataset are recorded at two spatial resolutions: 64 channels (actiChamp) and 32 channels (Brainamp MR). To ensure consistent input dimensionality, all recordings are mapped to a unified 31-channel representation. Let the raw signal be **X** ∈ ℝ ^*T* ×*C*^, where *T* denotes time steps and *C* the original channel count. The alignment mapping **X**_31_ is defined as:

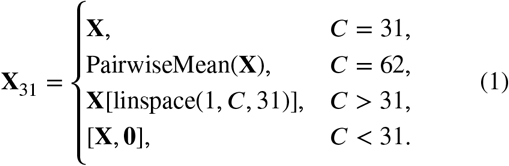

Specifically: for *C* = 62, PairwiseMean(**X**) averages pairs of physically adjacent channels in the high-density cap, preserving local spatial information while halving the channel count; for *C* > 31 (*C* ≠ 62), linearly spaced indexing selects 31 channels with uniform scalp coverage; for *C* < 31, zero-padding fills the missing channel dimensions.

EEG signals are highly susceptible to high-amplitude transient artifacts (e.g., eye blinks, muscle contractions) that can severely distort standard Z-score normalization. We therefore adopt a robust normalization scheme based on the median and median absolute deviation (MAD), applied independently to each channel’s time series. For the *j*-th channel *x*_*j*_ in **X**_31_:

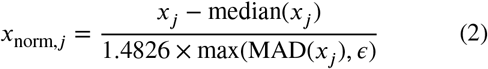

where MAD(*x*_*j*_) = median(|*x*_*j*_ −median(*x*_*j*_)|). The constant 1.4826 renders the estimator asymptotically equivalent to the standard deviation under Gaussian noise, and ϵ = 10^−6^ prevents division-by-zero in flat signal segments.

Deep neural networks require fixed-length inputs; feeding variable-length sequences directly would cause gradient instability and excessive memory consumption. We therefore introduce a dynamic overlapping sliding-window mechanism that segments each recording into fixed-length windows of *W* = 1024 time steps.

For recordings with *T* < 1024, zero-padding is applied at the end to reach the standard length. For *T* > 1024, *N* overlapping windows are extracted via linear equidistant sampling. The starting index *S*_*k*_ of the *k*-th window (*k* ∈ {0, 1, …, *N* − 1}) is:

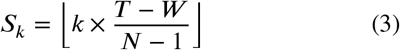

The *k*-th sub-signal is 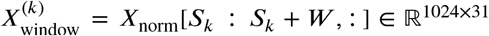 . To maximize data utilization, the number of windows *N* is set adaptively: *N*_train_ = 32 during training (providing implicit temporal augmentation) and *N*_val_ = 16 during validation (ensuring evaluation efficiency). Each segment is transposed to ℝ^31×1024^ before being fed into the CNN module.

### 3.2. Network Architecture

NSSI-related EEG signals are characterized by the coexistence of local rhythmic abnormalities (e.g., prefrontal Theta over-activation) and macroscopic long-range temporal dependencies (e.g., sustained frontolimbic coupling changes). To jointly capture these multi-scale properties, we design a cascaded 1D-CNN–BiGRU–MHSA architecture that achieves hierarchical mapping from local spatiotemporal micro-features to macroscopic temporal dynamics, enabling end-to-end NSSI risk prediction.

#### 3.2.1. Spatial-Temporal Micro-feature Extraction via 1D-CNN

To capture local microscopic waveform features associated with NSSI-related prefrontal abnormalities—such as excessive Theta-band activation or localized Gamma oscillations—the model’s front end deploys a spatiotemporal feature extractor comprising four cascaded one-dimensional convolutional blocks (ConvBlocks). The preprocessed EEG segment **X**_window_ ∈ ℝ^*N*×31^ (where *N* = 1024 time steps) is first transposed to ℝ^31×*N*^ and fed into the CNN. Each ConvBlock applies a uniform kernel size *K* = 31 and output channel count *F* = 64, maintaining consistent receptive field coverage across all four layers. The internal structure of each ConvBlock follows the sequence: Conv1d → BatchNorm1d → ReLU → MaxPool → Dropout.

For the *l*-th ConvBlock, given the input feature map **H**^(*l*−1)^, the output is:

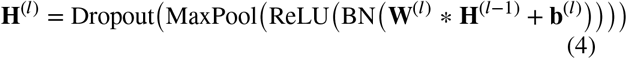

where * denotes the 1D convolution operation, BN(⋅) is batch normalization, and each MaxPool operation halves the temporal dimension. After four successive ConvBlocks, the time dimension is compressed from 1024 to 64 steps (1024 → 512 → 256 → 128 → 64), yielding an output of shape ℝ^64×64^ (64 time steps, 64 feature channels). This output is then transposed to ℝ^64×64^ as the sequential input to the BiGRU module, effectively encoding compact spatiotemporal micro-features while suppressing high-frequency background noise.

#### 3.2.2. Long-term Dynamic Modeling via BiGRU

The neural mechanisms underlying emotion dysregulation in NSSI patients often manifest as dynamic decay and reorganization of brain network connectivity states (e.g., prefrontal–amygdala coupling) on a timescale of hundreds of milliseconds. To capture these temporal evolutionary dynamics, the 64-step feature sequence output by the CNN module is fed into a Bidirectional Gated Recurrent Unit (BiGRU). Compared to standard RNNs, GRUs effectively mitigate the gradient vanishing problem inherent in long EEG sequences through learnable gating mechanisms.

In the forward GRU unit, for the input vector **x**_*t*_ ∈ ℝ^64^ at time step *t*, the update gate *z*_*t*_ and reset gate *r*_*t*_ are computed as:

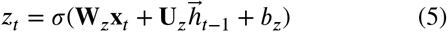

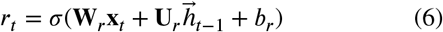

where *σ*(⋅) is the Sigmoid activation function, and **W, U** are learnable weight matrices. The reset gate *r*_*t*_ controls the degree to which past hidden state information is discarded, while the update gate *z*_*t*_ governs the proportion of the candidate state 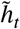 incorporated into the final hidden state. The candidate state and the forward hidden state 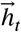 are updated as:

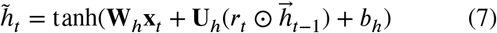

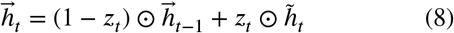

The backward GRU processes the reversed input sequence to compute 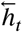. Each unidirectional hidden layer has dimension 256; the forward and backward states are concatenated to yield a 512-dimensional context-aware representation: 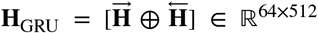, which encodes both past and future temporal context for each of the 64 time steps.

#### 3.2.3. Pathological Focus via Multi-Head Self-Attention

Not all EEG time segments contribute equally to NSSI diagnosis. To enable the model to adaptively focus on key temporal windows that reflect limbic system dysfunction, a multi-head self-attention (MHSA) layer with *H* = 8 heads is embedded after the BiGRU module.

The BiGRU output **H**_GRU_ ∈ ℝ^64×512^ is linearly projected into query (*Q*), key (*K*), and value (*V*) subspaces for each head. For the *i*-th attention head, the projections and scaled dot-product attention with dropout are:

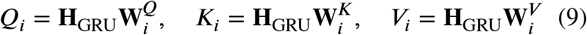

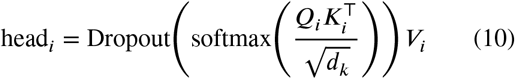

where *d*_*k*_ = 512/*H* = 64 is the per-head key dimension, and the scaling factor 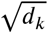 prevents excessively large dot products from causing Softmax saturation. The outputs of all *H* heads are concatenated and passed through a linear projection layer **W**^*O*^, followed by a residual connection and layer normalization:

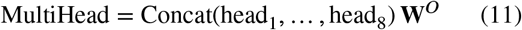

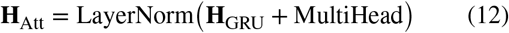

The residual connection and layer normalization ensure stable gradient flow during backpropagation. The resulting attention feature map **H**_Att_ ∈ ℝ^64×512^ encodes pathology-weighted temporal representations, with higher attention weights assigned to EEG segments most indicative of NSSI-related neural abnormalities.

#### 3.2.4. Global Aggregation and Classification Head

At the network’s output stage, global average pooling (*P*_avg_) and global max pooling (*P*_max_) are applied in parallel along the time axis of **H**_Att_, capturing complementary holistic representations:

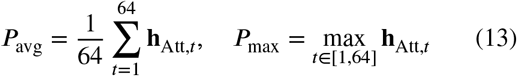

The two pooled vectors are concatenated to form a 1024-dimensional global descriptor **v** = [*P*_avg_ ⊕ *P*_max_] ∈ ℝ^1024^, which is then passed through a two-layer classification head consisting of a linear projection, dropout regularization, a second linear layer, and a Sigmoid activation:

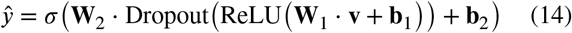

where **W**_1_, **b**_1_ and **W**_2_, **b**_2_ are the weights and biases of the first and second linear layers, respectively. The scalar output *ŷ* ∈ [0, 1] represents the posterior probability that the input EEG segment belongs to the NSSI class.

### 3.3. Loss Function

Clinical NSSI datasets are inherently class-imbalanced, with control samples substantially outnumbering NSSI cases. Standard binary cross-entropy (BCE) loss is susceptible to being dominated by the majority class during training.

We therefore adopt Focal Loss [32], which down-weights easy-to-classify samples via a dynamic modulating factor, directing optimization toward hard, near-boundary examples.

Let *p*_*t*_ denote the model’s predicted probability for the ground-truth class:

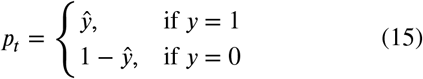

where *ŷ* is the model’s output probability and *y* ∈ {0, 1} is the ground-truth label. The Focal Loss is:

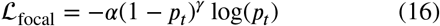

where *α* = 0.75 is the class-balancing weight and *γ* = 2.0 is the focusing parameter. The modulating factor (1 − *p*_*t*_)^*γ*^ exponentially down-weights high-confidence samples, concentrating optimization on hard examples near the decision boundary—such as early-stage NSSI patients with subtle EEG abnormalities.

To further smooth the decision boundary and improve generalization, we incorporate Mixup augmentation [33], applied with 50% probability per batch. Mixup linearly interpolates pairs of training samples and their labels, yielding soft targets. The total loss ℒ_total_ is decomposed accordingly:

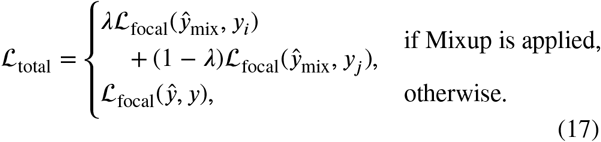

where *ŷ*_mix_ is the prediction for the mixed sample, and *y*_*i*_, *y*_*j*_ are the original labels of the two constituent samples. This joint loss design simultaneously addresses class imbalance and broadens the decision boundary, improving robustness to the complex variability of clinical EEG data.

## 4. Experiments

### 4.1. Datasets

Data were sourced from a project on cognitive neural mechanisms and biomarkers of graded emotional problems in children and adolescents conducted at the Shanghai Mental Health Center. The study protocol was approved by the institutional ethics committee via expedited review (Ethics No.: 2022-74C3), and written informed consent was obtained from all participants and their guardians. EEG signals were acquired using two device types: a 64-channel actiChamp system and a 32-channel Brainamp MR system. A total of 217 adolescents were enrolled and divided into an NSSI group (189 cases) and a non-NSSI control group (28 cases) based on the presence or absence of self-injurious behavior. Given the pronounced class imbalance, data were partitioned using three-fold cross-validation. Participant demographics are summarized in Table 1.

**Table 1.** Demographic characteristics of study participants.

| Group | n | Male/Female | Age (mean ± SD) | BMI (mean ± SD) | Only child (1/2) | Parental marital status (1/2/3) |
| --- | --- | --- | --- | --- | --- | --- |
| NSSI | 189 | 27/162 | 14.35 ± 1.60 | 21.76 ± 5.31 | 115/74 | 154/26/9 |
| Non-NSSI | 28 | 6/22 | 14.49 ± 1.32 | 21.62 ± 5.66 | 19/9 | 26/2/0 |
| Total | 217 | 33/184 | 14.37 ± 1.57 | 21.74 ± 5.30 | 134/83 | 180/28/9 |

### 4.2. Evaluation Protocol

Model performance is evaluated using five complementary metrics: accuracy (ACC), area under the ROC curve (AUC), specificity (SPE), sensitivity (SEN), and F1-score. These metrics are defined as:

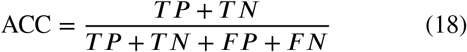

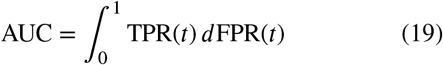

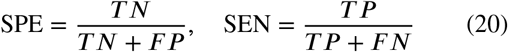

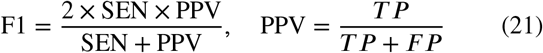

where *T P, T N, F P*, and *F N* denote true positives, true negatives, false positives, and false negatives, respectively. AUC measures discrimination ability across all classification thresholds; values approaching 1 indicate superior performance.

### 4.3. Implementation Details

All raw EEG recordings were first subjected to standardized offline preprocessing [29]: channel selection, bandpass filtering (0.5–45 Hz), notch filtering (50 Hz), resampling, and average referencing, converting .set files to .npy format. Subsequent processing followed the pipeline in Section 3.1. The framework was implemented in Python 3.10 with PyTorch, and all experiments were conducted on an NVIDIA GeForce RTX 5080 GPU. Training used the AdamW optimizer [34] with an initial learning rate and weight decay of 3 × 10^−5^. The learning rate schedule employed cosine annealing with warm restarts [35] (*T*_0_ = 30, *T*_mult_ = 2, minimum 1×10^−7^), preceded by a 15-epoch linear warmup from 5% of the target rate. Models were trained for up to 200 epochs with a batch size of 4; early stopping with patience of 50 epochs was applied to prevent overfitting.

### 4.4. Comparison Experiments

To validate the effectiveness of the proposed CGA-NSSI model in EEG-based identification of adolescent NSSI, we selected multiple representative models for comparison, including traditional machine learning methods (Logistic Regression, KNN [36], Random Forest [37]), general deep learning models (MLP [38], 1D-CNN [39], LSTM [40], Transformer [41]), and the NSSI-specific model NSSI-Net [30]. All models were trained and tested using the same data splitting strategy. Quantitative results are summarized in Table 2, and the corresponding visual comparison is shown in Fig. 2.

**Table 2.** Comparison of classification performance across methods (mean ± std). The proposed method is highlighted in bold.

| Method Type | Method | ACC | AUC | SPE | SEN | F1 |
| --- | --- | --- | --- | --- | --- | --- |
| Machine Learning | Logistic Regression | 0.461 ± 0.072 | 0.524 ± 0.053 | 0.781 ± 0.117 | 0.413 ± 0.097 | 0.567 ± 0.087 |
| | KNN [36] | 0.714 ± 0.187 | 0.525 ± 0.024 | 0.293 ± 0.236 | 0.778 ± 0.252 | 0.810 ± 0.155 |
| | Random Forest [37] | 0.797 ± 0.057 | 0.511 ± 0.036 | 0.359 ± 0.076 | 0.862 ± 0.080 | 0.880 ± 0.039 |
| Deep Learning | MLP [38] | 0.455 ± 0.164 | 0.480 ± 0.046 | 0.685 ± 0.195 | 0.423 ± 0.216 | 0.554 ± 0.197 |
| | 1D-CNN [39] | 0.493 ± 0.128 | 0.589 ± 0.101 | 0.859 ± 0.051 | 0.439 ± 0.151 | 0.591 ± 0.144 |
| | LSTM [40] | 0.525 ± 0.130 | 0.603 ± 0.053 | 0.793 ± 0.200 | 0.487 ± 0.175 | 0.627 ± 0.158 |
| | Transformer [41] | 0.553 ± 0.024 | 0.590 ± 0.062 | 0.715 ± 0.057 | 0.529 ± 0.033 | 0.673 ± 0.024 |
| | NSSI-Net [30] | 0.644 ± 0.067 | 0.679 ± 0.071 | 0.776 ± 0.080 | 0.654 ± 0.061 | 0.725 ± 0.040 |
|  | **CGA-NSSI (Ours)** | **0.708 ± 0.038** | **0.742 ± 0.102** | **0.819 ± 0.100** | **0.735 ± 0.033** | **0.762 ± 0.031** |

**Figure 2.**
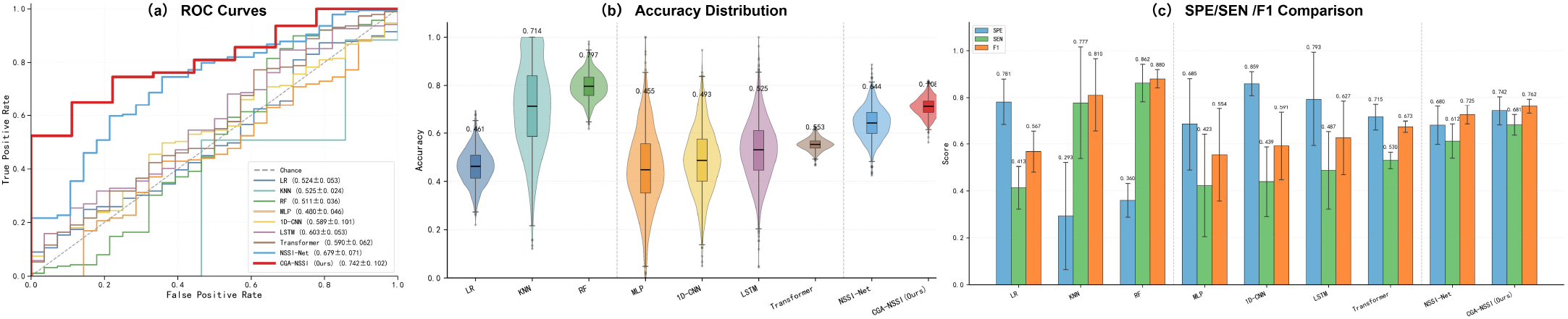
Performance comparison across methods. (a) ROC curves showing CGA-NSSI consistently above all baselines. (b) ACC comparison bar chart. (c) F1-score violin plot illustrating distribution stability.

Traditional machine learning methods perform relatively weakly overall. Logistic Regression achieves an AUC of only 0.524, reflecting the inability of linear classifiers to capture the complex nonlinear structure of NSSI EEG. KNN and Random Forest improve ACC and F1 but their AUCs (0.525 and 0.511) remain limited, indicating poor class-level discrimination, consistent with the summary in Fig. 2.

General deep learning models show greater potential: 1D-CNN raises AUC to 0.589 by extracting local temporal patterns; LSTM reaches 0.603 by modeling sequential dependencies; Transformer achieves 0.590 via self-attention over long-range context. However, since NSSI EEG simultaneously exhibits local rhythmic structure and long-range dynamic dependencies, single-architecture models cannot fully exploit multi-level pathological information. The NSSI-specific NSSI-Net achieves ACC/AUC/F1 of 0.644/0.679/0.725, representing a meaningful step forward, yet its overall discriminative ability remains below that of our model, as also reflected by the ROC, ACC, and F1 visualizations in Fig. 2.

CGA-NSSI achieves the best results across all metrics: ACC = 0.708 ± 0.038, AUC = 0.742 ± 0.102, SPE = 0.819 ± 0.100, SEN = 0.735 ± 0.033, F1 = 0.762 ± 0.031. Relative to NSSI-Net, CGA-NSSI improves AUC, ACC, and F1 by approximately 9.3%, 9.9%, and 5.1%, respectively. The cascaded 1D-CNN–BiGRU–MHSA design simultaneously captures local spatiotemporal micro-features and long-range temporal dynamics, with attention mechanisms reinforcing the representation of pathologically salient segments. The balanced SPE and SEN further confirm the model’s suitability for clinical screening, where both false negatives (missed NSSI cases) and false positives (unnecessary referrals) carry significant consequences.

### 4.5. Ablation Study

To validate the necessity and synergistic contribution of the three core modules (1D-CNN, Bi-GRU, and multi-head self-attention) in CGA-NSSI, we conducted ablation experiments by progressively removing key components. Results are shown in Table 3.

**Table 3.** Ablation study results (mean ± std). ✓ indicates that the module is included, and the full CGA-NSSI model is highlighted in bold.

| 1D-CNN | BiGRU | Attention | ACC | AUC | SPE | SEN | F1 |
| --- | --- | --- | --- | --- | --- | --- | --- |
| ✓ |  |  | 0.493 ± 0.128 | 0.589 ± 0.101 | 0.859 ± 0.051 | 0.439 ± 0.151 | 0.591 ± 0.144 |
|  | ✓ |  | 0.715 ± 0.360 | 0.587 ± 0.030 | 0.500 ± 0.045 | 0.804 ± 0.075 | 0.804 ± 0.044 |
| ✓ | ✓ |  | 0.617 ± 0.224 | 0.605 ± 0.121 | 0.711 ± 0.192 | 0.603 ± 0.277 | 0.691 ± 0.237 |
| ✓ |  | ✓ | 0.695 ± 0.138 | 0.637 ± 0.006 | 0.648 ± 0.172 | 0.704 ± 0.184 | 0.717 ± 0.121 |
|  | ✓ | ✓ | 0.723 ± 0.121 | 0.643 ± 0.087 | 0.644 ± 0.096 | 0.715 ± 0.143 | 0.815 ± 0.087 |
| ✓ | ✓ | ✓ | **0.708 ± 0.038** | **0.742 ± 0.102** | **0.819 ± 0.100** | **0.735 ± 0.033** | **0.762 ± 0.031** |

With only 1D-CNN, the model extracts local temporal features (AUC = 0.589, SPE = 0.859) but achieves SEN of only 0.439, reflecting insufficient temporal modeling. With only BiGRU, dynamic dependencies are captured (ACC = 0.715, SEN = 0.804), but the absence of local feature extraction yields AUC = 0.587 and SPE = 0.500. Two-module combinations show progressive improvement: 1D-CNN + BiGRU reaches AUC = 0.605; 1D-CNN + Attention improves to 0.637; BiGRU + Attention achieves ACC = 0.723 and F1 = 0.815 but AUC = 0.643. Only the full three-module CGA-NSSI achieves the best performance across all metrics (AUC = 0.742) with the smallest standard deviations, confirming that each component addresses a distinct and complementary aspect of the NSSI EEG classification problem.

### 4.6. Electrode Importance Analysis

To investigate the spatial distribution of neural abnormalities in resting-state EEG of NSSI individuals and to identify key EEG channels for precise NSSI recognition, we employed the Integrated Gradients method with positive-negative attribution difference to quantify the discriminative contribution of each electrode to the inter-group classification task. Combined with scalp topographic maps, we analyzed the spatial distribution differences in EEG signals between the two groups, and validated the classification effectiveness of key electrode subsets of different sizes.

As shown in Fig. 3 and Fig. 4, electrode importance analysis results indicate that the central midline electrode Cz exhibits significantly higher inter-group discriminative contribution than other electrodes, serving as the core channel for distinguishing NSSI individuals from healthy controls. Electrodes Fp1, O2, P3, CPz, TP7, and Iz show secondary discriminative contributions. High-importance electrodes are not uniformly distributed across the scalp but show significant spatial clustering, primarily covering the central integration region, left prefrontal control region, and occipitoparietal posterior representation region. This suggests that NSSI-related EEG abnormalities are not focal changes in a single brain region, but rather a cross-regional spatial organizational shift along the central-posterior-prefrontal axis.

**Figure 3.**
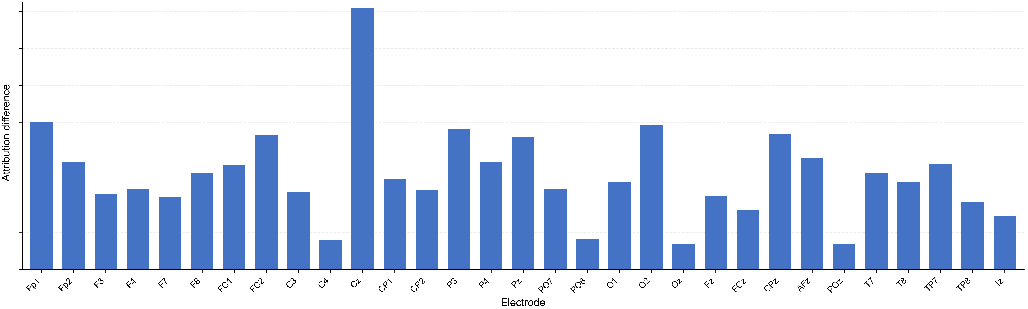
Electrode-wise model attribution for self-harm prediction under the 10–20 EEG system. Bar height denotes each electrode’s predictive contribution, with Cz exhibiting the dominant attribution weight.

**Figure 4.**
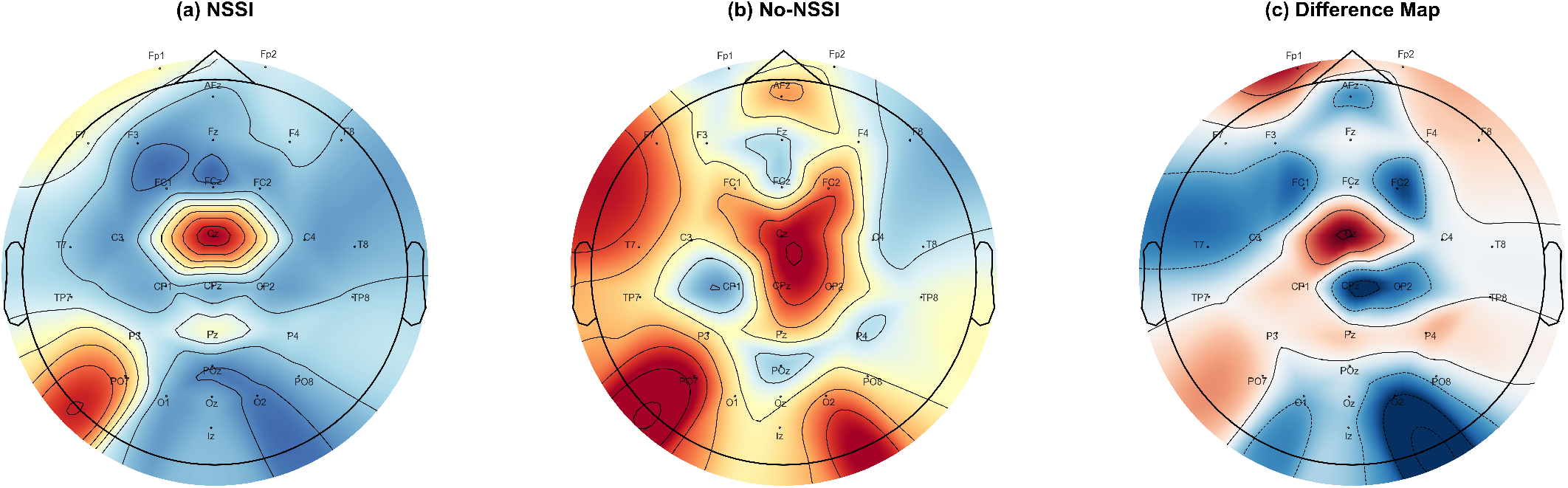
Scalp topographic plots of predictive attribution values for the NSSI group (a), non-NSSI control group (b), and their intergroup difference map (c). Warm colors denote high attribution weight while cool colors represent low contribution across scalp electrodes.

As shown in Table 4, the lightweight discriminative model built based on electrode importance ranking shows an overall increasing trend in classification performance as the number of selected key channels increases. The Top-15 electrode subset achieves the best discriminative performance, with accuracy, AUC, and F1-score of 0.728 ± 0.017, 0.634 ± 0.047, and 0.830 ± 0.014, respectively, approaching the performance of the full-channel CGA-NSSI model. This confirms that the core electrodes within this subset can carry the primary neural feature information for NSSI recognition.

**Table 4.** Classification performance of electrode subsets of varying sizes.

| Channels | ACC | AUC | SPE | SEN | F1 |
| --- | --- | --- | --- | --- | --- |
| Top-3 | 0.478 ± 0.260 | 0.571 ± 0.095 | 0.659 ± 0.337 | 0.455 ± 0.349 | 0.523 ± 0.307 |
| Top-5 | 0.561 ± 0.256 | 0.535 ± 0.094 | 0.548 ± 0.335 | 0.566 ± 0.343 | 0.617 ± 0.316 |
| Top-10 | 0.552 ± 0.214 | 0.545 ± 0.085 | 0.537 ± 0.364 | 0.556 ± 0.292 | 0.628 ± 0.267 |
| Top-15 | 0.728 ± 0.017 | 0.634 ± 0.047 | 0.500 ± 0.045 | 0.762 ± 0.026 | 0.830 ± 0.014 |
| All 31 | 0.708 ± 0.038 | 0.742 ± 0.102 | 0.819 ± 0.100 | 0.735 ± 0.033 | 0.762 ± 0.030 |

In summary, the central midline, left prefrontal, and occipitoparietal posterior regions are the core spatial areas of resting-state EEG abnormalities in NSSI individuals, with cross-regional neural organizational reconstruction as the key pathological feature. The key electrode subset identified in this study provides a potential reference scheme for efficient, lightweight EEG screening of NSSI.

### 4.7. Brain Region Difference Analysis

Building on the spatial feature conclusions from electrode importance and scalp topographic distribution analyses, this section further analyzes the neural connectivity abnormality patterns in NSSI individuals at the brain network level. Based on EEG functional connectivity, we construct inter-group difference networks between the NSSI group and healthy controls, quantifying functional connectivity mean, inter-group difference, effect size, and statistical significance across five core brain regions: frontal, occipital, central, temporal, and parietal lobes.

As shown in Fig. 6, the inter-group difference functional connectivity network is primarily concentrated in the frontal, central, and occipital regions, with dense abnormal functional connections between brain regions; temporal region connection differences are secondary, while parietal inter-group abnormal connections are relatively sparse. Regional root mean square (RMS) difference results clearly indicate that the frontal lobe shows the highest inter-group connection difference amplitude, followed by occipital, central, and temporal regions in decreasing order, with the parietal lobe showing the lowest overall difference amplitude. This clearly identifies the frontal-central-occipital axis as the core spatial range of NSSI functional connectivity abnormalities, which is also consistent with the regional summary shown in Fig. 5.

**Figure 5:**
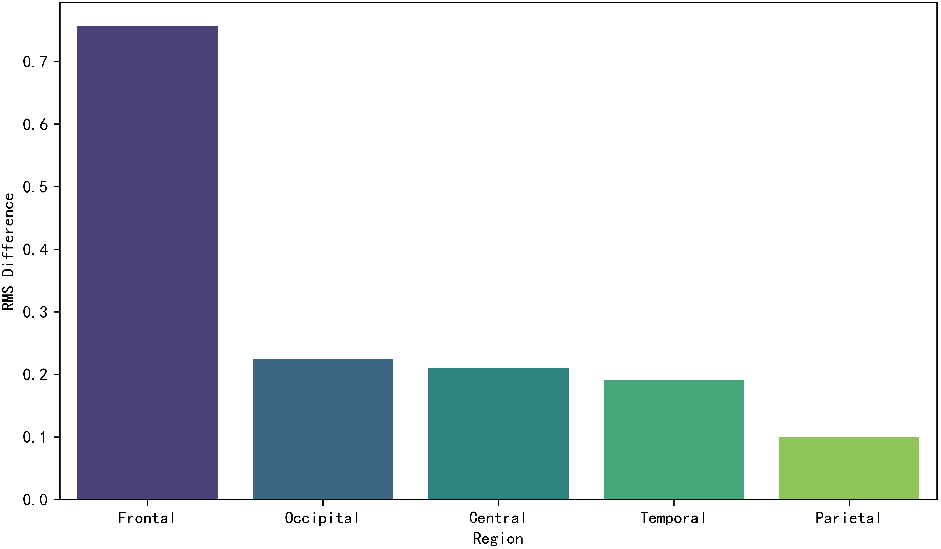
Regional RMS difference across five brain lobes. Frontal lobe exhibits the largest intergroup divergence, followed by occipital, central, temporal and parietal regions.

**Figure 6.**
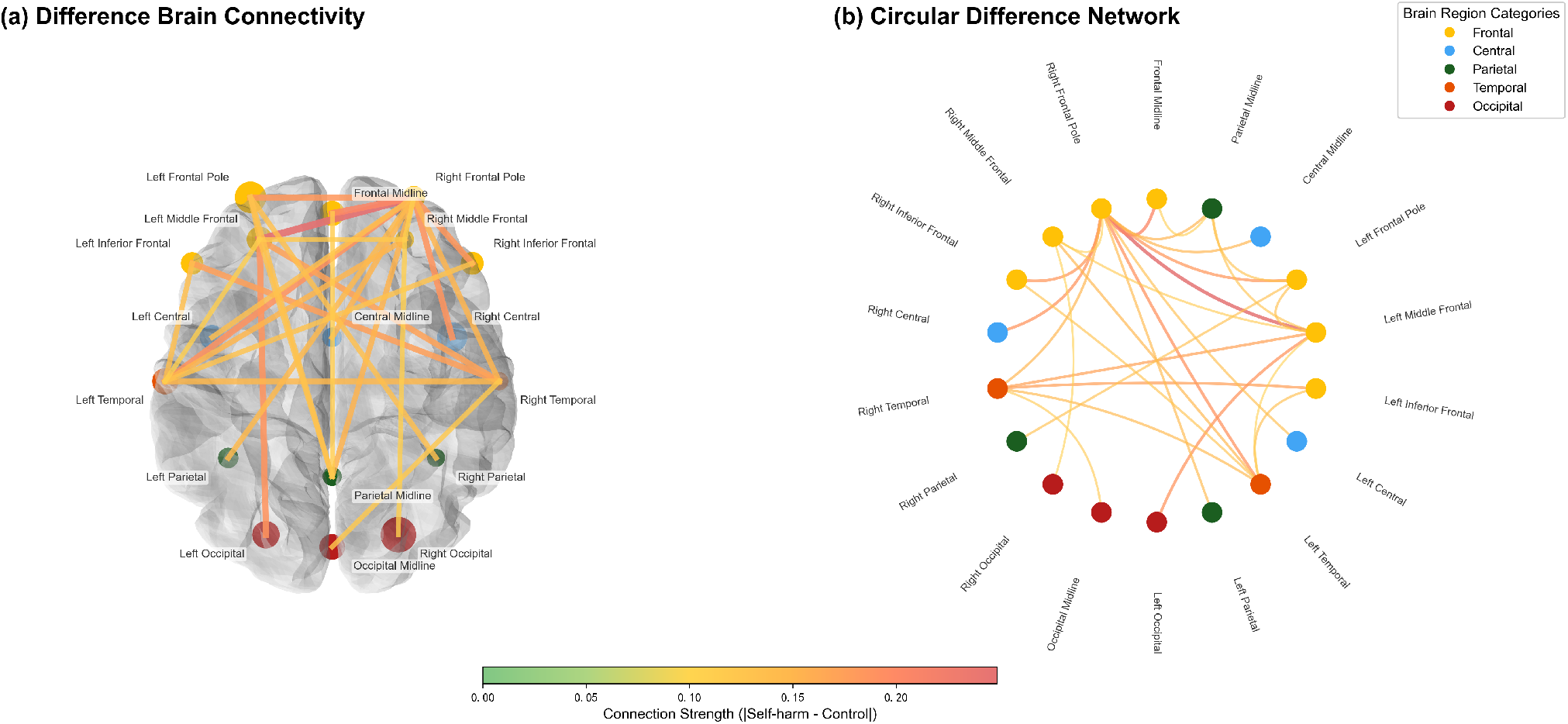
Brain region difference analysis. (a) Inter-group difference functional connectivity network. (b) Regional RMS difference bar chart. The frontal-central-occipital axis shows the highest inter-group differences.

Combined with regional quantitative statistical results (Table 5), the functional connectivity mean of all five brain regions in the NSSI group is higher than in the healthy control group. The frontal lobe shows the highest inter-group difference (Diff = 0.756) and effect size (Cohen’s *d* = 0.182) across the whole brain, with occipital, central, and temporal regions showing decreasing differences and effect sizes, and the parietal lobe showing the weakest overall inter-group change. Statistical results show that the parietal lobe has a relatively smaller *p*-value (*p* = 0.027), while other brain regions have *p*-values greater than 0.05. However, considering the combined difference, effect size, and network visualization results, the parietal lobe shows only minor inter-group changes and is not the core brain region of NSSI-related neural abnormalities. Overall, NSSI individuals primarily exhibit enhanced functional connectivity in the frontal-central-occipital axis.

**Table 5.** Inter-group functional connectivity statistics across five brain regions. Diff = NSSI mean − Control mean.

| Region | NSSI | Control | Diff | Cohen's *d* | *p* |
| --- | --- | --- | --- | --- | --- |
| Frontal | 2.513 | 1.757 | 0.756 | 0.182 | 0.082 |
| Occipital | 2.219 | 1.995 | 0.224 | 0.073 | 0.106 |
| Central | 1.883 | 1.673 | 0.211 | 0.076 | 0.077 |
| Temporal | 1.955 | 1.764 | 0.191 | 0.066 | 0.117 |
| Parietal | 1.926 | 1.826 | 0.100 | 0.035 | 0.027 |

In summary, the brain region functional connectivity difference analysis results are highly consistent with the core conclusion of cross-regional spatial organizational shift along the central-posterior-prefrontal axis obtained in the previous section, further confirming at the network level that NSSI-related EEG abnormalities are not focal changes in a single brain region, but rather a network reorganization involving coordinated imbalance across multiple brain regions.

### 4.8. Static Functional Connectivity Analysis

Building on the multi-region network imbalance features revealed in the brain region analysis, this section employs a static functional connectivity (sFC) analysis framework to further investigate the neural network abnormality patterns in resting-state EEG of NSSI individuals from three perspectives: whole-brain neural synchronization patterns, core abnormal connectivity pathways, and the discriminative value of global connectivity levels.

As shown in Fig. 7, the whole-brain static functional connectivity matrix provides a direct visual comparison of neural synchronization differences between the two groups. Compared to healthy controls, the NSSI group shows overall higher functional connectivity strength, with whole-brain neural synchronization showing an overall upward trend; the inter-group difference matrix is dominated by positive differences, indicating a widespread enhancement of signal coupling between brain regions in the self-injury group. This type of connectivity level increase does not represent positive optimization of brain function, but rather reflects pathological features of brain network regulatory imbalance and redundant cross-regional signal coupling.

**Figure 7.**
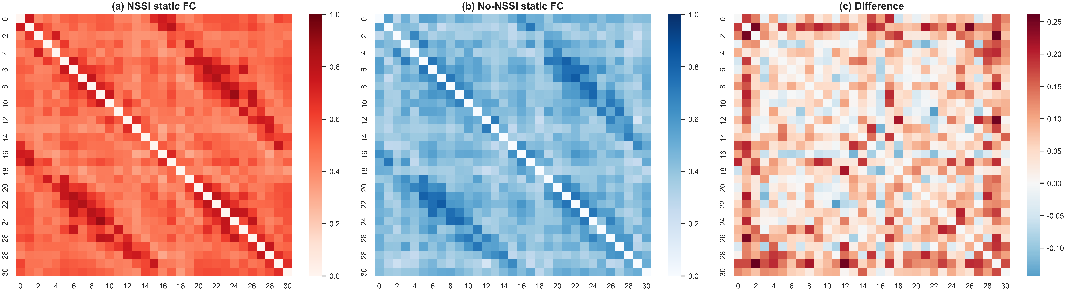
Static functional connectivity matrices for the NSSI group (a), non-NSSI controls (b), and their intergroup difference map (c). Warm colors denote elevated connectivity strength.

As shown in Fig. 8, extracting the top 20 functional connectivity pathways with the largest inter-group difference amplitudes reveals that core abnormal connections are highly concentrated in cross-regional coupling pathways between the frontal-temporal and frontal-occipitoparietal regions. As shown in Table 6, the 5 most prominent connections all show statistically significant differences (*p* < 0.05), primarily involving frontal-temporal and frontal-occipitoparietal pathways such as F3↔TP8, CP2↔TP8, and Fp2↔F3. The coupling abnormalities between the frontal lobe’s core emotion regulation regions and the temporal lobe’s emotion processing and occipitoparietal posterior perceptual integration regions are most pronounced. This result is highly consistent with the central-posterior-prefrontal spatial abnormality pattern confirmed earlier, again confirming the central role of the frontal lobe hub in NSSI neural abnormalities.

**Table 6.** Top 5 functional connectivity edges with the largest inter-group differences (all *p* < 0.05).

| Edge | NSSI | Control | Diff | $p$ |
| --- | --- | --- | --- | --- |
| F3 ↔ TP8 | 0.473 | 0.211 | 0.261 | 0.010 |
| CP2 ↔ TP8 | 0.426 | 0.173 | 0.253 | 0.014 |
| Fp2 ↔ F3 | 0.665 | 0.416 | 0.249 | 0.016 |
| Fp2 ↔ T7 | 0.604 | 0.357 | 0.247 | 0.015 |
| C3 ↔ TP8 | 0.517 | 0.302 | 0.215 | 0.025 |

**Figure 8.**
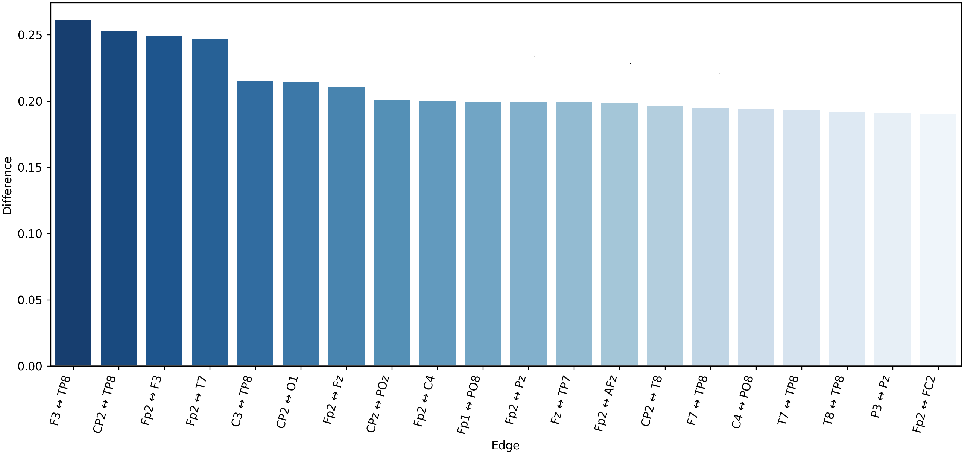
Ranked bar plot showing intergroup difference of static functional connectivity across top altered EEG electrode pairs, with bar height representing connectivity divergence between NSSI patients and healthy controls.

To investigate the intrinsic relationship between static functional connectivity physiological features and deep learning model discriminative results, a window-level correlation analysis was conducted using a sliding window strategy. EEG signals were segmented with a window size of 1024 time points and a step size of 512 time points; the Pearson correlation coefficient matrix of 31 electrode channels within each window was computed, with the upper triangular matrix mean representing the global mean of whole-brain static functional connectivity for that window. Each window signal was then input to the trained model to obtain the corresponding NSSI discrimination probability. Spearman correlation analysis across approximately 62,000 window samples showed a weak but significant positive correlation between the global mean of whole-brain static functional connectivity and model discrimination probability (*ρ* = 0.101, *p* = 0.00826, *q* = 0.0124), with statistical results remaining robust after multiple testing correction. This result indicates that higher overall brain synchronization is associated with a higher probability of the model classifying a sample as NSSI, validating that global functional connectivity upregulation is an important physiological basis for model NSSI recognition, as illustrated in Fig. 9.

**Figure 9.**
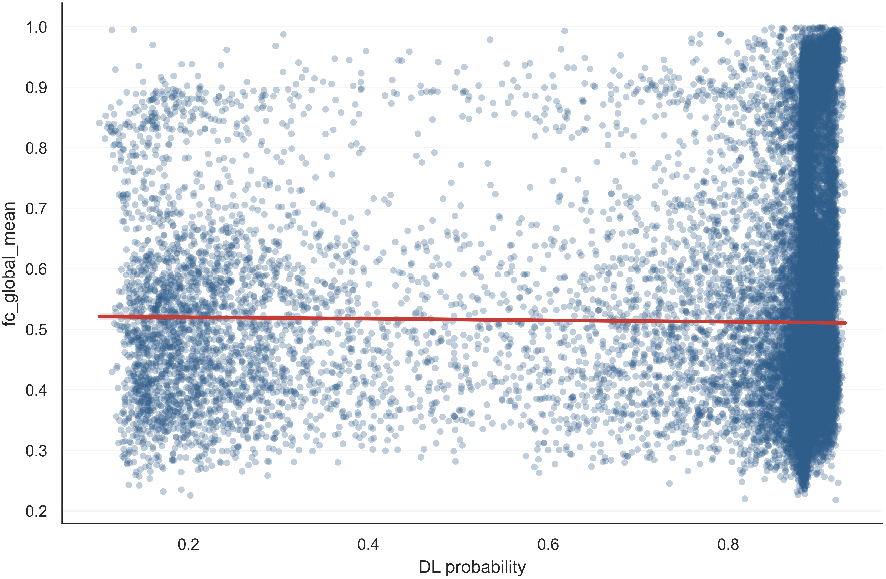
Scatter plot illustrating the correlation between global mean functional connectivity and deep learning derived NSSI risk probability. The red horizontal line denotes the population averaged global connectivity threshold.

In summary, resting-state EEG in NSSI individuals shows overall upregulation of whole-brain neural synchronization, with core abnormalities concentrated in the frontal-temporal-occipitoparietal pathway over-coupling, and global connectivity levels can effectively reflect self-injury risk. These results, combined with electrode importance and brain region difference analyses, form a complete evidence chain, further elucidating the neural circuit mechanisms underlying impaired emotion regulation and impulse control in NSSI individuals.

### 4.9. Dynamic Functional Connectivity Analysis

Building on the abnormal features of whole-brain neural synchronization upregulation and cross-regional signal over-coupling revealed in the static functional connectivity analysis, this section extends the research perspective to dynamic functional connectivity (DFC), investigating the dynamic abnormality patterns of neural regulation in NSSI individuals from the perspective of temporal evolution of brain networks. Combined with interpretable machine learning, risk stratification analysis, and clinical behavioral scale data, we explore the intrinsic relationship between dynamic neural features and self-injury-related psychological behaviors.

Starting with core dynamic indicators of brain networks for inter-group comparison (Fig. 10), we selected switching rate, transfer entropy, and fluctuation degree to measure the switching frequency, switching disorder degree, and temporal fluctuation amplitude of whole-brain connectivity, respectively. Results show that switching rate and transfer entropy do not exhibit significant inter-group differences, but overall show a trend of higher values in the self-injury group; fluctuation degree shows a clear statistical difference (*p* = 0.038), with the healthy control group showing higher temporal fluctuation amplitude of brain network. This result indicates that the brain network dynamic flexibility of self-injury individuals is weaker, with an overall more rigid neural regulation pattern, making it difficult to achieve flexible temporal response and regulation.

**Figure 10.**
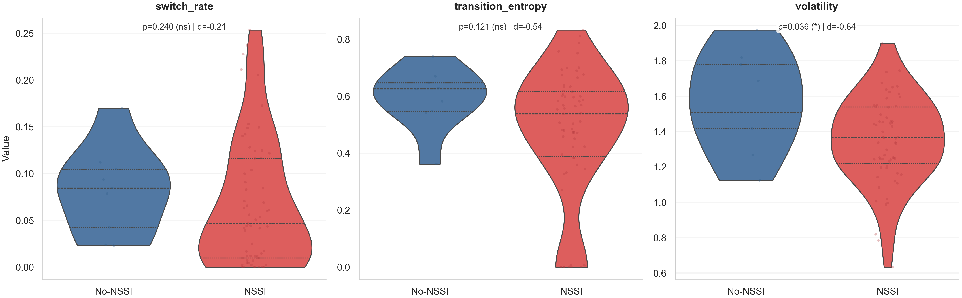
Violin plots comparing core dynamic functional connectivity metrics between control and self harm groups. The self harm group exhibits significantly lower volatility.

Further K-means clustering analysis stably divided the dynamic brain network into two core patterns with clear physiological and behavioral significance (Fig. 11, Table 7). State 0 is a low-connectivity recovery state with a global average connectivity strength of 0.436, where brain regions operate relatively independently, representing the brain’s baseline stable standby recovery mode. State 1 is a high-connectivity emotional occupation state with a global average connectivity strength of 0.786, where the brain network is highly synchronously activated, corresponding to a stress state where emotional resources are heavily occupied and cognitive control ability is reduced. From state occupancy rate and average dwell time, healthy controls maintain the normal recovery state for the vast majority of time (occupancy rate 91.6%), entering the emotional activation state only rarely; while the NSSI group’s dwell proportion in the high-connectivity emotional state significantly increases to 23.6%, and once entering this state, the average dwell window is as long as 58.3, far exceeding the control group’s 2.0, directly reflecting that self-injury individuals’ brains are more prone to falling into abnormally coupled emotional networks and difficult to exit autonomously.

**Table 7.** Dynamic functional connectivity states and their functional interpretations.

| Metric | State 0 (Low-connectivity) | State 1 (High-connectivity) |
| --- | --- | --- |
| Global edge mean ± SD | 0.436 ± 0.080 | 0.786 ± 0.075 |
| Fractional occupancy – Self-harm | 76.4% | 23.6% |
| Fractional occupancy – Control | 91.6% | 8.4% |
| Mean dwell time – Self-harm (windows) | 113.5 | 58.3 |
| Mean dwell time – Control (windows) | 33.2 | 2.0 |
| Functional interpretation | Resting / recovery state | Emotion-occupied / stress-activated state |

**Figure 11.**
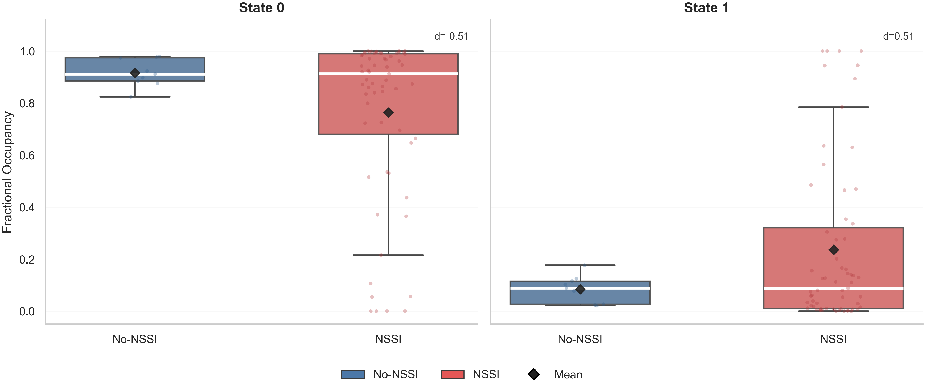
Boxplots of fractional occupancy for State 0 and State 1 comparing control and self harm groups. Black diamonds denote group means.

To validate the clinical behavioral significance of the two network states, correlation analysis with multi-dimensional psychological scales was conducted. The detailed associations are summarized in Table 8. Results show a complementary correspondence pattern between State 0 and State 1. Greater occupancy or longer dwell time in State 0 tends to align with the opposite direction of the same scales linked to State 1, consistent with the interpretation of State 0 as a recovery / low-stress state. By contrast, longer dwell time or greater occupancy in State 1 tends to co-occur with poorer peer relationship, higher internet addiction, higher perceived stress, and more sleep problems, supporting its interpretation as a high-connectivity emotional occupation state. Because these associations did not survive FDR correction, they should be regarded as exploratory behavioral support rather than confirmatory evidence.

**Table 8.** Clinical scale correspondence for State 0 and State 1 dynamic functional connectivity metrics.

| State | DFC Metric | Scale | Spearman $\rho$ | p-value |
| --- | --- | --- | --- | --- |
| State 0 | Fractional occupancy | Sleep problems | +0.250 | 0.061 |
| State 0 | Fractional occupancy | Internet addiction | +0.216 | 0.107 |
| State 0 | Fractional occupancy | Peer relationship | +0.195 | 0.142 |
| State 0 | Mean dwell time | Sleep problems | +0.284 | 0.032 |
| State 0 | Mean dwell time | Childhood trauma | +0.169 | 0.210 |
| State 0 | Mean dwell time | Internet addiction | +0.164 | 0.224 |
| State 1 | Fractional occupancy | Sleep problems | -0.250 | 0.061 |
| State 1 | Fractional occupancy | Internet addiction | -0.216 | 0.107 |
| State 1 | Fractional occupancy | Peer relationship | -0.195 | 0.142 |
| State 1 | Mean dwell time | Peer relationship | -0.329 | 0.012 |
| State 1 | Mean dwell time | Internet addiction | -0.269 | 0.043 |
| State 1 | Mean dwell time | Perceived stress | -0.258 | 0.053 |

Note: State 0 and State 1 fractional occupancy are complementary, so their correlations appear with opposite signs. These associations are exploratory and do not survive FDR correction.

The state transition matrix further reveals differences in network switching logic between the two groups (Fig. 12). Healthy individuals have good self-regulation ability and can quickly return to normal mode even after briefly entering an abnormal network state; while self-injury individuals, once trapped in an abnormal network, show significantly reduced probability of switching back to the normal state, making it difficult to autonomously recover to neural homeostasis.

**Figure 12.**
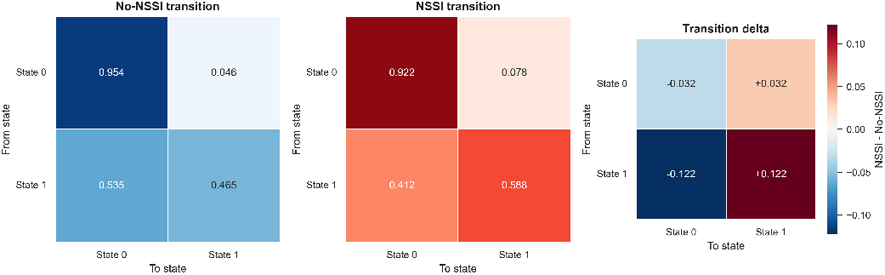
State transition matrices for control, self-harm groups and their difference. Color bar shows transition probability differences.

SHAP interpretability analysis quantified the contribution weights of various dynamic indicators to model recognition results (Fig. 13). Network state transitions, state occupancy rates, and network switching-related indicators are the key dynamic features for the model to accurately identify self-injury individuals, with abnormal state-related indicators and network switching features showing particularly prominent positive contributions, confirming the close relationship between dynamic neural abnormalities and self-injury phenotype at the algorithmic level.

**Figure 13.**
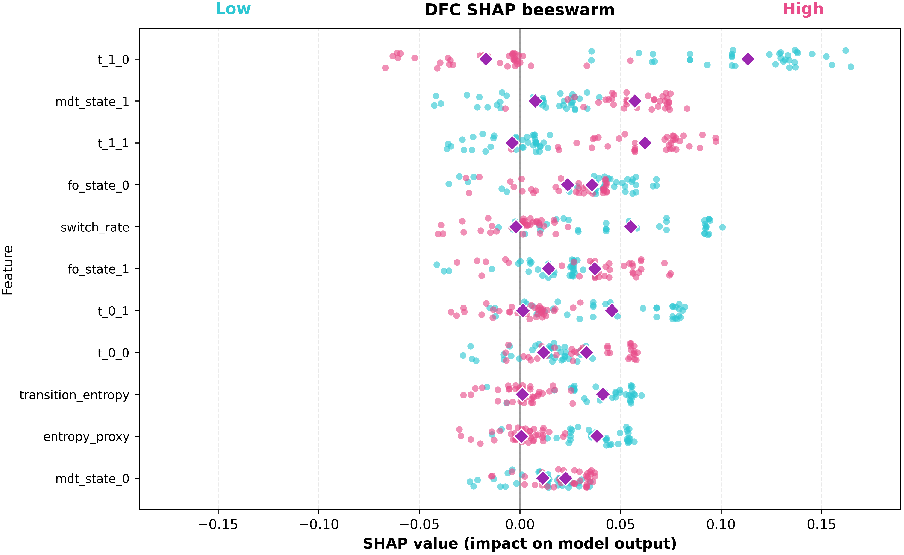
SHAP beeswarm plot for DFC features in self harm classification using random forest. Each point shows a sample’s SHAP value, colored by feature magnitude (low cyan to high pink), illustrating feature impact on model output.

Based on the deep learning model’s discrimination probability, samples were divided into low, medium, and high self-injury risk stratification levels to explore the gradient change patterns of dynamic indicators with increasing risk (Fig. 14). Results show that as the self-injury risk level increases, the occupancy proportion of the normal network state gradually decreases, the dwell probability in the abnormal state continuously increases, and the dynamic switching pattern of the brain network also undergoes pathological shifts synchronously, confirming that the degree of dynamic functional connectivity abnormality is positively correlated with self-injury risk.

**Figure 14.**
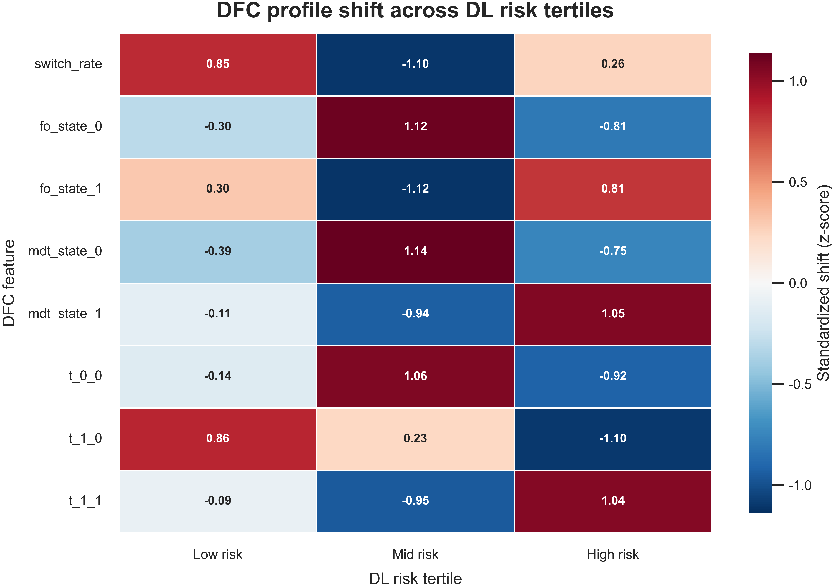
Heatmap of standardized DFC feature shifts across DL risk tertiles. Color intensity reflects z-score magnitude.

Overall, the neural abnormalities in NSSI individuals are not only reflected at the static level of whole-brain over-coupling, but also manifest as temporal regulatory imbalance at the dynamic level, with core manifestations of reduced brain network flexibility, tendency to fall into high-synchronization emotional activation states, and insufficient self-recovery ability. This section combines traditional dynamic brain network analysis, clinical behavioral correlation, interpretable deep learning, and risk stratification validation, forming a complete closed loop with the electrode importance, brain region difference, and static functional connectivity analyses, jointly elucidating the neural circuit mechanisms underlying impaired emotion regulation and impulse control in self-injury individuals from spatial-temporal-behavioral multiple dimensions.

## 5. Discussion

This study exploits the high temporal resolution of EEG to both validate the efficacy of CGA-NSSI in self-injury risk screening and systematically reveal the neuroelectrophysiological mechanisms underlying adolescent NSSI. Through Integrated Gradients attribution, SHAP interpretability, and multi-dimensional functional connectivity analysis (sFC/dFC), we integrate resting-state spatial network reorganization with dynamic temporal evolution features, fusing deep learning-derived quantitative indicators with clinical psychological phenotypes to construct a complete spatial–temporal–behavioral pathological evidence chain.

At the static spatial and brain network level, the NSSI group exhibits significant overall upregulation of whole-brain neural synchronization, with this abnormality highly concentrated in the frontal lobe internally and in its connections with the central region. This connectivity enhancement does not represent positive optimization of brain function, but rather reflects pathological over-coupling of the prefrontal cortex and compensatory dysregulation of cognitive control. Facing intense negative emotions, adolescent patients may continuously exhaust prefrontal resources for emotional suppression, leading to loss of local network flexibility and significantly reduced cognitive reappraisal efficiency; when this compensatory mechanism collapses under extreme emotional pressure, it easily transforms into self-injurious impulses. Further edge tracing reveals that the most significantly different pathways (e.g., F3-TP8, CP2-TP8) broadly involve cross-regional communication between the prefrontal cortex and the temporoparietal junction and central posterior regions. This suggests that NSSI neural abnormalities are not limited to the emotional network, but extend to systemic integration disorders spanning advanced cognitive control, somatosensory processing, and self-awareness. When the cognitive system’s modulation of the temporoparietal-somatosensory pathway is severely imbalanced, individuals are more inclined to counteract vague and intolerable psychological pain by disrupting bodily boundaries and introducing definite physical pain, consistent with the somatization defense mechanism whereby patients report using physical pain to override intolerable psychological distress.

At the dynamic temporal level, DFC analysis further characterizes the rigidity of neural dynamics in NSSI individuals. Results show that the brain network temporal fluctuation degree in the self-injury group is significantly lower, and they are more readily trapped in the highly synchronized high-connectivity emotional state (State 1) during state transitions. This decline in brain functional state switching ability perfectly maps to the clinical phenotype of rumination in patients, the subjective experience of emotional entrapment and the objective failure of neural state-switching are two sides of the same pathological coin. Furthermore, SHAP attribution analysis confirms that the transition feature from the high-connectivity stress state to the low-connectivity baseline state is the key dynamic indicator for the model to accurately identify self-injury risk. This reveals the core dynamic pathology of self-injurious behavior: impaired ability to return to baseline. Since the neural system’s capacity to self-reset from an abnormally high arousal state is substantially impaired, patients are forced to use external strong stimuli (such as the strong pain input from cutting) to forcibly interrupt the infinite loop of negative emotions, achieving a destructive form of self-regulation.

In summary, four interrelated pathological features identified in this study—prefrontal compensatory over-coupling, frontal-temporoparietal integration disorder, neural dynamics rigidity, and impaired baseline recovery—together constitute a comprehensive neuropathological hypothesis for adolescent NSSI. These findings not only provide solid neurophysiological evidence for the black-box decisions of the deep learning model, but also provide important scientific references for future selection of non-invasive neural intervention targets and early clinical psychological counseling for self-injurious adolescents.

Several limitations warrant acknowledgment. First, data collection relied on clinically presenting adolescents, resulting in a pronounced class imbalance (189 NSSI vs. 28 controls) that may constrain generalization to community-based populations. Second, this work is restricted to the EEG modality; future studies should incorporate structural and functional neuroimaging to provide a more comprehensive mechanistic account of adolescent NSSI.

## 6. Conclusion

The neurophysiological complexity of adolescent NSSI and the inherent imbalance of clinical EEG data present formidable barriers to early precise screening. Our work addresses both challenges through the lightweight CGA-NSSI framework, which not only achieves state-of-the-art classification performance across all metrics but also demonstrates the decisive role of multi-level spatiotemporal feature mining and key electrode selection in capturing subtle EEG abnormalities. Beyond diagnostic accuracy, CGA-NSSI delivers neurobiological interpretability by revealing prefrontal compensatory over-coupling and impaired neural state-switching as core pathological signatures of adolescent NSSI. Future work will incorporate larger, balanced cohorts and explore multi-modal data fusion to advance this early-warning model toward real-world clinical deployment and deeper mechanistic understanding.

## Data Availability

The authors do not have permission to share data.

## Data availability

The authors do not have permission to share data.

## Acknowledgments

This work was supported by grants from STI 2030— Major Projects (no. 2022ZD0209100); Natural Science Foundation of China (no. 82571771) and Natural Science Foundation of Shanghai (no. 25ZR1401167).

